# A statistical model for forecasting probabilistic epidemic bands for dengue cases in Brazil

**DOI:** 10.1101/2025.06.12.25329525

**Authors:** Laís Picinini Freitas, Danielle Andreza da Cruz Ferreira, Raquel Martins Lana, Daniel Cardoso Portela Câmara, Tatiana P. Portella, Marilia Sá Carvalho, Ayrton Sena Gouveia, Iasmim Ferreira de Almeida, Eduardo Correa Araujo, Luã Bida Vacaro, Fabiana Ganem, Oswaldo Gonçalves Cruz, Flávio Codeço Coelho, Claudia Torres Codeço, Luiz Max Carvalho, Leonardo Soares Bastos

## Abstract

Dengue is a vector-borne disease and a major public health concern in Brazil. Its continuing and rising burden has led the Brazilian Ministry of Health to request for modelling efforts to aid in the preparedness and response to the disease. In this context, we propose a Bayesian forecasting model based on historical data to predict the number of cases 52 weeks ahead for the 118 health districts of Brazil. We leverage the predictions to build probabilistic epidemics bands to be used for dengue monitoring. We define four disjoint probabilistic bands (≤50%, (50%,75%], (75%,90%], and ≥90%) based on the percentiles of the predicted cases distribution and interpreted according to the historical number of cases and past occurrence probability (below the median, typical; moderately high, fairly typical; fairly high, atypical; exceptionally high, very atypical). We performed out-of-sample validation for 2022-2023 and 2023-2024 and forecasted 2024-2025. In the 2022-2023 and 2023-2024 seasons, the epidemic bands followed the observed cases’ curve shape, with a sharp increase after January and a decline after the peak around April. In 2022-2023, the observed number of cases (1,436,034) was slightly above the estimated 75% percentile (1,405,191), being classified as “fairly high, atypical”. Most health districts in South Brazil showed exceptionally high numbers of cases during this season. The situation worsened in 2023-2024 and the observed number of cases (6,454,020) was way above the 90% percentile (2,221,557), characterising an “exceptionally high, very atypical” season. For the 2024-2025 season, we estimated a median number of cases of 1,526,523 (maximum value for the “below the median, typical” probabilistic epidemic band. The maximum estimated values for the upper bands were 2,213,282 (moderately high, fairly typical) and 3,803,898 (fairly high, atypical) with the upper limits of the probabilistic epidemic bands of 1,452,359. Probabilistic epidemic bands serve as a valuable monitoring tool by enabling prospective comparisons between observed case curves and historical epidemic patterns, facilitating the assessment of ongoing outbreaks about past occurrences.

## 1. Introduction

Dengue is a vector-borne disease on the rise globally caused by four different viral serotypes and transmitted to humans by *Aedes aegypti* and *Aedes albopictus* mosquitoes. In the Americas only, dengue affected around 13 million people and caused more than 8 thousand deaths in 2024, exceeding the region’s historical annual record (PAHO, 2025; Lenharo, 2023). In the same year, Brazil faced its most severe dengue epidemic since the introduction of the DENV-1 and DENV-4 serotypes in Roraima in 1981 (Osanai et al., 1983). Over 6.6 million cases and 6 thousand related deaths were reported (Brasil, Ministério da Saúde, 2025). This dire situation has motivated the Brazilian Ministry of Health to reach out for modelling approaches that could guide interventions in a timely manner.

Forecasting models can be useful for enhancing public health preparedness by anticipating potential epidemic outbreaks and informing resources allocation and timely interventions, especially when they provide estimates in high spatial and temporal resolutions. Specifically for dengue, forecasting models have been proposed using statistical methodologies with different levels of complexity (Colón-González et al., 2021b; Pirani et al., 2024; Karasinghe et al., 2024) and, in recent years, machine learning methods (Stolerman et al., 2019; Al Mobin, 2024; Sebastianelli et al., 2024). With different methodologies presenting interesting results for both explaining and predicting dengue outbreaks, an important question remains on what methods can be accurate while being useful and operationally viable for health authorities.

In this paper, we propose a Bayesian spatio-temporal forecasting model based on historical surveillance data to predict dengue cases up to 52 weeks in the future for all 118 health districts of Brazil. These predictions correspond to what would be expected under similar conditions as those observed in the past. Therefore, they can be used for monitoring to detect atypical scenarios. To develop this monitoring tool, we build probabilistic epidemic bands based on percentiles of the predictive cases distributions and interpret them considering the historical median and the probability of past occurrence.

## 2. Methods

### 2.1. Study context and data

A health district is an administrative grouping of neighbouring municipalities from the same Federal Unit (state) that share socioeconomic and transport infrastructure (Supplementary Figure A.1). Their purpose in the Brazilian Unified Health System (*Sistema Único de Saúde* - SUS) is to organise the health care network in an integrated manner to correct access inequalities.

We used surveillance data from the Notifiable Diseases Information System (*Sistema de Informação de Agravos de Notificação* - SINAN) from the Brazilian Ministry of Health, where dengue is a disease of mandatory reporting. A reported case of dengue corresponds to a patient who sought medical care and received a suspected or confirmed diagnosis of dengue upon medical evaluation. Later, a reported case can be discarded for reasons such as confirmation of another aetiological agent.

Anonymised dengue case data are openly available at the individual level from https://datasus.saude.gov.br/transferencia-de-arquivos/. Cases missing information on the municipality of residence were excluded. The municipality of residence was used to identify the health district of residence of cases. Most of dengue cases occur in summer and fall, so we opt to start a season in spring, on epidemiological week 41, and ending a season on week 40 of the following year. Then, cases were grouped by health district of residence, season and season week of symptoms onset. We included notified cases of dengue symptoms onset between October 11, 2015, and October 05, 2024, and excluded all discarded cases, obtaining a dataset of “probable cases”.

### 2.2. Statistical model

We propose a Bayesian negative binomial model with structured random effects for season week and unstructured random effects for the season for each health district. The forecast was performed retrospectively for the seasons 2022-2023 and 2023-2024 and prospectively for season 2024-2025. Predictions of the number of dengue cases are obtained from samples of an approximate posterior predictive distribution per week and health district. These predictions can be further aggregated to lower spatial and temporal resolutions (e.g. Federal Units and season).

#### 2.2.1. Model specification

Let *Y_t,h_*be the number of dengue cases at time *t* and health district *h*. We then assume that

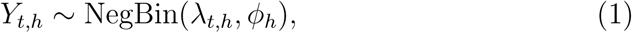

where *ϕ_h_ >* 0 is an overdispersion parameter and *λ_t,h_* is the expected number of dengue cases at time *t* in health district *h*, modelled as

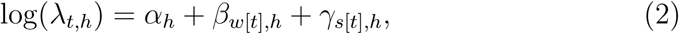

where *α_h_* are health district-specific intercepts, *β_w_*_[*t*]*,h*_ are cyclic second-order random walk effects by week *w*, *w* = 1, 2*, … ,* 52, and health district *h*, and *γ_s_*_[*t*]*,h*_ independent Gaussian random effects for season *s*, *s* = 1, 2*, … , S*, and health district *h*, *h* = 1, 2*, … , H*. The cyclic term on the week random effects means that the first week, week 1, is correlated with the last one, week 52.

The model specification is completed by setting penalized complexity (PC) priors for the remaining parameters (Simpson et al., 2017). For the overdispersion parameter *ϕ_h_* a PC Gamma is chosen with a limiting Poisson case as a baseline. For the random effects variances 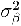 and 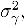 the same PC prior for the precision with parameters (3,0.01) such that *P* (*σ >* 3) = 0.01, where the standard deviation *σ* = *τ* ^1*/*2^.

The posterior predictive distribution of the cases of the following year, *s* = *S* + 1, for each health district, *h*, and all 52 weeks is given by

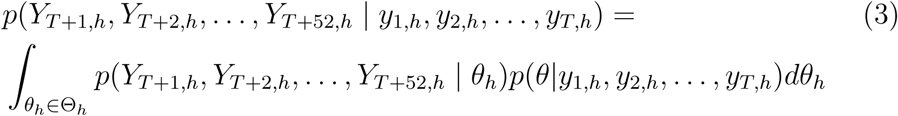

where 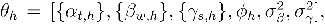 the first term inside the integral is the likelihood (1) evaluated at the new data and the second is the posterior distribution of *θ_h_*. Inference is done via integrated nested Laplace approximation (Rue et al., 2009; Martins et al., 2013), and samples from an approximation for the posterior predictive distribution (3) using the Gaussian latent parameters and the discrete version of the hyperparameters distribution can be obtained the inla.posterior.sample function implemented in the INLA package.

#### 2.2.2. Probabilistic epidemic bands and classification

From the samples, we obtain the 50th, 75th, and 90th percentiles of the predictive distribution of the cases for each health district, *h*, and week, *w*. These percentiles establish probabilistic epidemic thresholds, delineating four distinct probability bands of ≤50%, (50%,75%], (75%,90%], and ≥90%. We classify these probabilistic epidemic bands based on the historical case distribution and past occurrence probabilities, designating them as follows: below the median, typical; moderately high, fairly typical; fairly high, atypical; and exceptionally high, very atypical (Table 1). The historical median is defined as the 50th percentile of the predicted case distribution from 2015 up to the season immediately preceding the one being forecasted.

**Table 1:** Proposed classification of predicted cases level with the interpretation.

| Band | Predicted cases level | Interpretation |
| --- | --- | --- |
| $\leq 50\%$ | Below the median, typical | Number of cases below the historical median, with a 50% occurrence probability. |
| 50-75% | Moderately high, fairly typical | Number of cases moderately higher than the historical median, with a 25% occurrence probability. |
| 75-90% | Fairly high, atypical | Number of cases fairly higher than the historical median, with a 15% occurrence probability. |
| $\geq 90\%$ | Exceptionally high, very atypical | Number of cases exceptionally higher than the historical median, with a 10% occurrence probability. |

#### 2.2.3. Model validation

We performed out-of-sample validation for two seasons. For forecasting the 2022-2023 season, we used data by epidemiological week 40/2022, while for forecasting 2023-2024 we modelled data by epidemiological week 40/2023. We visually compared the predictive posterior probabilistic epidemic bands with the observed values of number of cases by season week at the national and Federal Unit levels. For each season, we classified and depicted the health districts according to the epidemic band within which the observed number of dengue cases fell.

Analyses were performed in R (version 4.4.1) (R Core Team, 2024) using R-INLA (version 24.06.27) (Rue et al., 2009; Martins et al., 2013). All data and code are available at https://github.com/AlertaDengue/baseline_ paper/.

## 3. Results

In this section, we first present the results of the retrospective forecasting for two seasons (2022-2023 and 2023-2024) and compare them with the observed number of dengue cases. Next, we show the results of the prospective forecasting for the 2024-2025 season.

### 3.1. Retrospective forecasting and model validation

For the 2022-2023 season, the observed number of dengue cases (1,436,034) was slightly above the estimated 75% percentile (1,405,191), falling within the 75-90% probabilistic epidemic band and classified as “fairly high, atypical” - with a low probability of occurrence considering the historical data (Table 2). The 2023-2024 season was very atypical, with an unprecedented number of observed cases of over 6.4 million. This number vastly exceeded the 90% percentile of the number of predicted cases, with a very low probability of occurrence.

**Table 2:** Percentiles of the predicted number of dengue cases by season and number of observed cases, 2022-2025, Brazil.

|  |  | 2022-2023 | 2023-2024 | 2024-2025 |
| --- | --- | --- | --- | --- |
| Percentile | 50% | 980,013 | 1,070,672 | 1,526,523 |
|  | 75% | 1,405,191 | 1,452,359 | 2,213,282 |
|  | 90% | 2,371,785 | 2,221,557 | 3,803,898 |
| Observed |  | 1,436,034 | 6,454,020 |  |

In Figure 1, the national-level weekly estimations are depicted against the curve of observed dengue cases for the 2022-2023 (panel A) and 2023-2024 (panel B) seasons. The colours depict the probabilistic epidemic bands, which indicate the predicted case level as proposed in Table 1. In both panels of Figure 1, we observe that the epidemic band shapes follow the curve of the observed cases, with a sharp increase after January and a decline after the peak around April.

**Figure 1:**
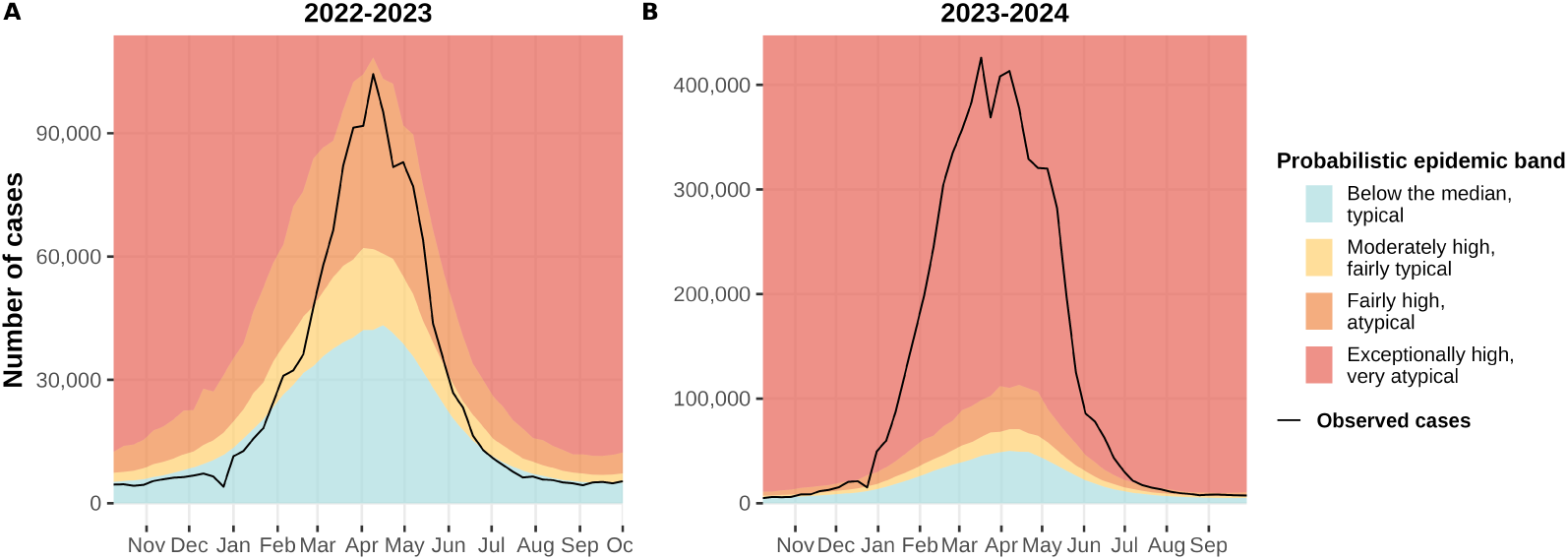
Estimated probabilistic epidemic bands compared with the observed number of dengue cases by week, 2022-2023 (A) and 2023-2024 (B) seasons, Brazil.

In 2022-2023 (Figure 1A), the observed cases curve fell within the “below the median, typical” band (in cyan) at the beginning of the season, quickly increasing and reaching the “moderately high, fairly typical” (in yellow) and “fairly high, atypical” (in orange) after February. Only during the peak (season week 27, in April), the observed number of cases was “exceptionally high, very atypical” (in red).

A markedly different scenario emerged in the 2023–2024 season, during which the observed dengue case curve remained within the “exceptionally high, very atypical” band for most of the season (Figure 1B). This band classification represents a highly unlikely occurrence based on historical data.

The probabilistic epidemic bands by week for each Federal Unit against the observed cases curve is depicted in Figure 2 for seasons 2022-2023 and 2023-2024.

**Figure 2:**
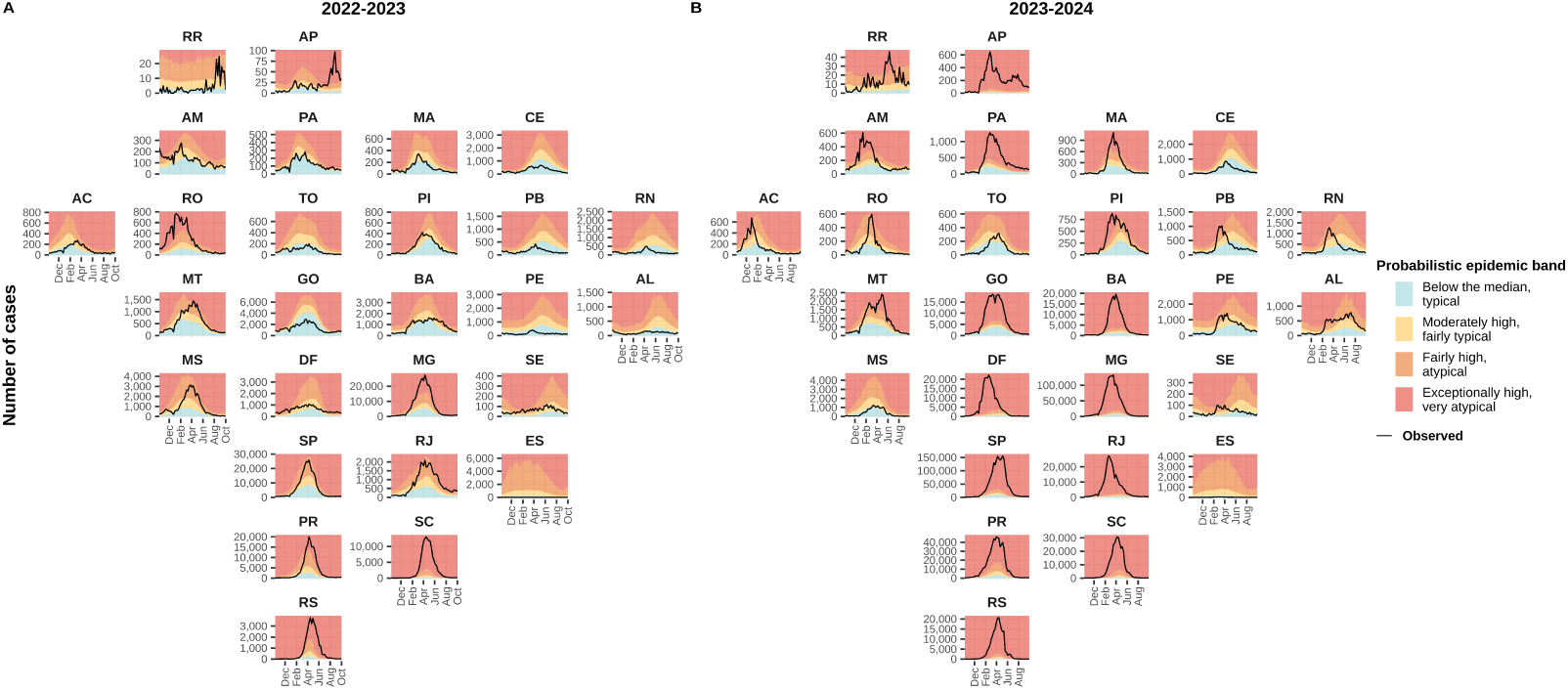
Estimated probabilistic epidemic bands compared with the observed number of dengue cases by week and Federal Unit in 2022-2023 (A) and 2023-2024 (B) seasons.

Many Federal Units had fewer observed cases than the historical median in 2022-2023 (Figure 2A), especially in Northeast Brazil (Maranhão - MA, Ceará - CE, Rio Grande do Norte - RN, Paraíba - PB, Pernambuco - PE, and Alagoas - AL). On the other hand, all southern Federal Units (Paraná - PR, Santa Catarina - SC, and Rio Grande do Sul - RS) had an exceptionally high number of dengue cases. This very atypical situation in the South continued during the 2023-2024 season (Figure 2B) when other Federal Units from all Brazilian regions also had an exceptionally high number of observed cases (North: Amapá - AP and Pará (PA); Mid-West: Goiás - GO and Federal District (DF); Northeast: Bahia - BA; Southeast: Minas Gerais - MG, São Paulo - SP, and Rio de Janeiro - RJ). Interestingly, for the 2023-2024 season, the curve of observed dengue cases in Paraíba (PB), Rio Grande do Norte (RN), Pernambuco (PE) and Alagoas (AL) peaked earlier than predicted considering the historical data.

The maps in Figure 3 illustrate the probabilistic epidemic band in which the total observed dengue cases for seasons 2022-2023 (panel A) and 2023-2024 (panel B) fell within across different health districts. By increasing the spatial resolution, we can clearly identify heterogeneity within Federal Units. For instance, during the 2022–2023 season, one health district in Amazonas (AM) experienced an exceptionally high number of dengue cases, while the rest of the state recorded case counts below the historical median. Although some health districts reported exceptionally high case numbers in 2022–2023 (30 out of 118), the majority had either fewer (45 out of 118) or moderately higher (27 out of 118) cases than the historical median (Figure 3A). In contrast, during the 2023-2024 season (Figure 3B), most health districts (70/118, or 59.3%) experienced an exceptionally high number of dengue cases, underscoring the unprecedented scale and severity of this epidemic.

**Figure 3:**
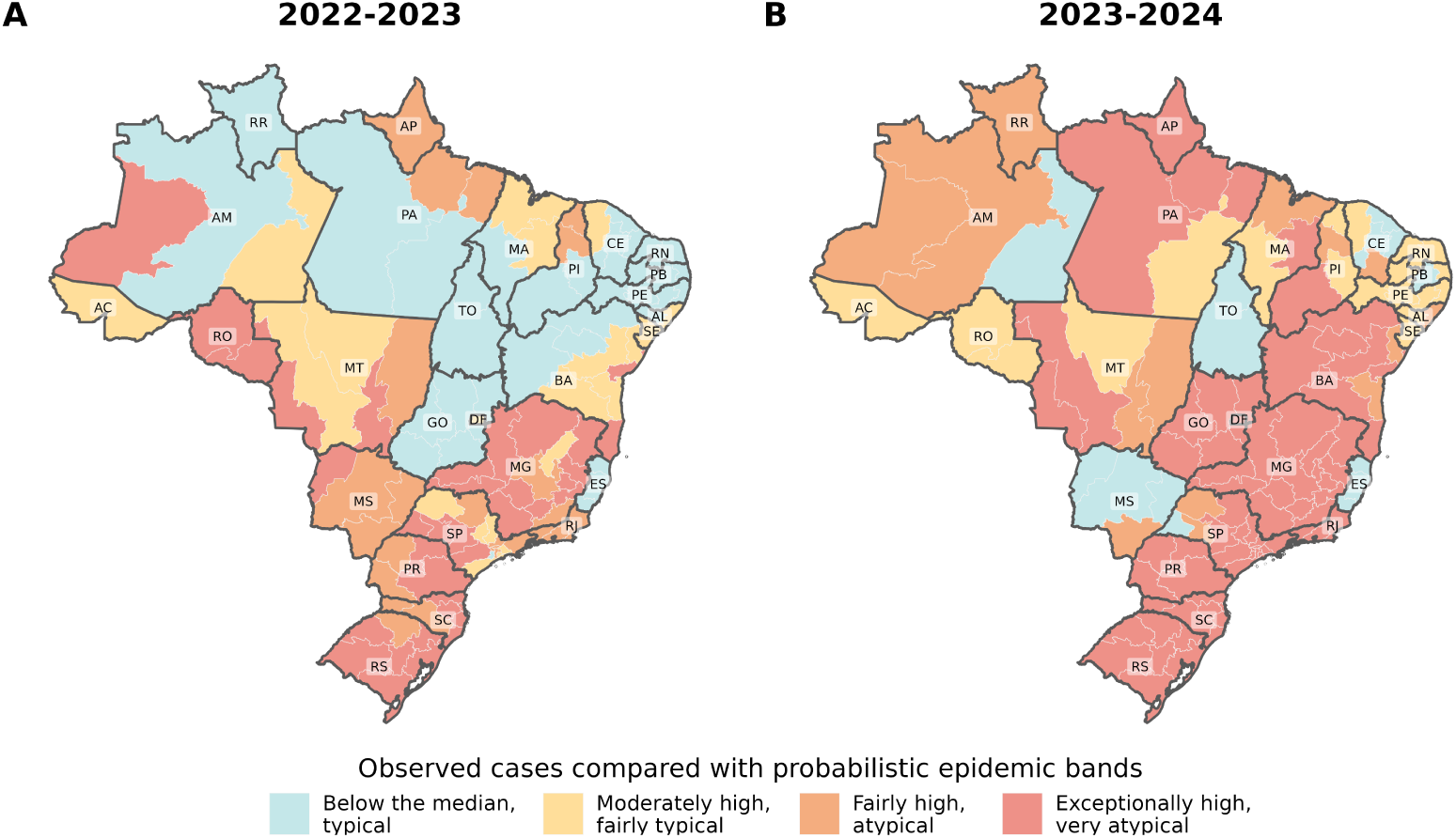
Observed number of cases compared with probabilistic epidemic bands by health district in the 2022-2023 (A) and 2023-2024 (B) seasons.

### 3.2. Prospective forecasting for the 2024-2025 season

For the 2024-2025 season, the estimated upper limits for the probabilistic epidemic bands were 1,526,523 (50%), 2,213,282 (75%) and 3,803,898 (90%) dengue cases in Brazil (Table 2). These represent a substantial increase in the predicted number of dengue cases compared to the estimates for the previous two seasons, influenced mainly by including the 2023-2024 season data in the model.

Figure 4A also shows the increase in the thresholds of the forecasted probabilistic bands of dengue cases for the 2024-2025 season compared to the previous two seasons. Figure 4B depicts the forecasted probabilistic epidemic bands for each Federal Unit. An increase in the thresholds is evident for MG, SP, PR, SC and RS, as well as for GO, RJ and BA. These Federal Units experienced an exceptionally high number of cases in 2023-2024.

**Figure 4:**
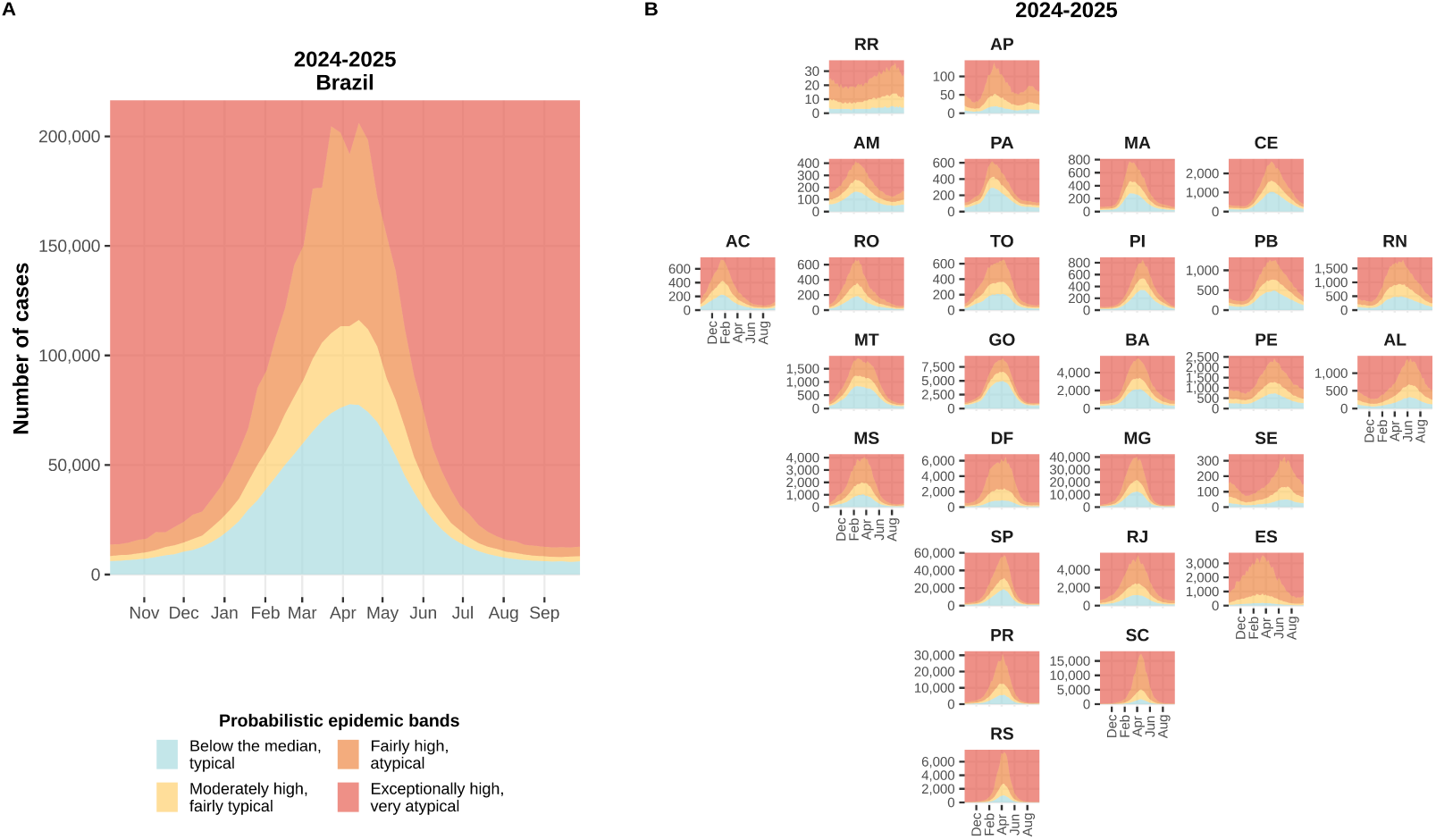
Forecasted probabilistic epidemic bands of weekly dengue cases for the 2024-2025 season in Brazil (A) and by Federal Unit (B).

## 4. Discussion

We successfully estimated probabilistic epidemic bands for dengue in Brazil using a forecasting model with only case number data. The predictions, derived from historical data, provide a baseline of what is expected for the next dengue season, with uncertainty, under conditions similar to those in the past. Due to this characteristic, our model predictions are helpful for prospective outbreak monitoring. To facilitate its use as such, we propose a classification of probabilistic epidemic bands that facilitates the visualization and interpretation of an ongoing dengue situation.

The out-of-sample validation demonstrated that our model generates reliable predictions for more typical dengue seasons. However, its accuracy declines in regions experiencing recent or ongoing dengue emergence (e.g., southern Brazil) or during seasons that deviate significantly from historical patterns (e.g., the 2023–2024 season). Since the model relies exclusively on historical case data, it cannot anticipate epidemic scenarios that have never occurred in the past. The 2023-2024 epidemic was unprecedented, more than 3 times bigger than the second-largest dengue epidemic in Brazil in 2018-2019 (Supplementary Figure A.2). This highly atypical season was likely the result of a combination of dengue expansion and intensified transmission dynamics. Dengue has been expanding geographically to new areas, putting previously unaffected populations at risk, a phenomenon that has intensified in recent years, likely due to global warming (Codeco et al., 2022; Barcellos et al., 2024). In addition, changes in meteorological conditions promoted by the El Niño Southern Oscillation (ENSO) climatic event may have favoured the biology of the vector *Ae. aegypti* and boosted both dengue expansion and transmission (Pirani et al., 2024; Vincenti-Gonzalez et al., 2018).

Therefore, climate, population immunity, and serotype circulation information could enhance our model when aiming at accurate and precise predictions. Data on population immunity and serotype circulation are often very limited. On the other hand, climate data have been increasingly made available and used to model vector-borne diseases (de Sousa et al., 2018; Colón-González et al., 2013, 2023; Sophia et al., 2025; Lowe et al., 2015, 2017; Chou-Chen et al., 2023; Leung et al., 2023; Colón-González et al., 2021b,a). We found that dengue outbreaks in Northeast Brazil occurred sooner than expected in 2023-2024. Suppose this was a consequence of changes in temperature or precipitation patterns. In that case, including climate data could have predicted the anticipation of the epidemic curve in the short term. In the long term, however, to estimate future climate data would be necessary. Additionally, the model would have to predict changes in climate patterns, which is not a trivial task. To address this challenge, long-term predictions with climate data may be built for different climatic scenarios (Colón-González et al., 2023, 2013; Sophia et al., 2025; Colón-González et al., 2021a). This, however, may compromise the usability of the predictions by the public health system. Such more complex structures considering climate factors, which may or may not have lagged and non-linear effects (Lowe et al., 2018; Ortega-Lenis et al., 2024; Lowe et al., 2021), can be explored in future work and by other research groups using our model as a “baseline model”.

As a monitoring tool, it is easy to detect early if a dengue situation differs from historical patterns with the probabilistic epidemic bands to generate alerts in a timely manner. This feature is further strengthened in combination with nowcasting models, which provides estimates of case numbers corrected for reporting delays in disease surveillance data (Bastos et al., 2019; McGough et al., 2020; Stoner and Economou, 2020; Günther et al., 2021). This aligns with the objectives of the “Action Plan for Reducing Dengue and other Arboviruses” from the Brazilian Ministry of Health(Brasil, Ministério da Saúde, 2024). In the “National Contingency Plan for Dengue, Chikungunya and Zika”, the Ministry of Health defines response strategies for four risk levels that can straightforwardly fit the four probabilistic bands we propose (Brasil, Ministério da Saúde et al., 2025). The plan presents a monitoring tool based on quantiles and medians of past distributions. In this regard, applying our model instead represents a similar idea with the same goal but an improvement for epidemic monitoring due to its statistical robustness.

This study has limitations regarding the nature of the data and the model. We used surveillance data, which is known to be prone to under-reporting. Under-reporting may vary in space-time, which hinders the ability of a model to estimate it. Laboratory confirmation is low among our study population, and other co-circulating diseases with similar clinical symptoms could be reported as dengue cases. As mentioned, dengue serotype circulation is an important factor influencing dengue epidemics. However, such information is limited, lacking enough spatio-temporal resolution, and is not publicly available in Brazil. Finally, our model does not account for population- and ecological-level factors that can contribute to dengue transmission and distribution. However, models based only on case data have the advantage of being more straightforward to implement and apply in different contexts. In a forecasting challenge for predicting neuroinvasive disease caused by West Nile virus, another mosquito-borne disease, simple models based on historical cases generally performed better than more complex models (Holcomb et al., 2023).

## 5. Conclusions

Brazil has continental dimensions and a large proportion of dengue control actions are coordinated at the national level. Therefore, identifying the heterogeneity in the magnitude and timing of the epidemic scenarios, both between and within Federal Units, is essential for better public health preparedness and response. Forecasting tools can assist in planning interventions, making them more precisely focused in both time and space. The fact that our forecasting model only needs case data makes it widely applicable. Both our model and the proposed classification of probabilistic epidemic bands can be applied in different countries and for other diseases, provided it is endemic and systematic surveillance data are available over a sufficient period. We recommend using at least five years of data and conducting out-of-sample validation to effectively assess the model’s performance.

## Data Availability

All data and code are available at https://github.com/AlertaDengue/baseline_paper/.

## Acknowledgements

We would like to thank the InfoDengue (https://info.dengue.mat.br/) and Mosqlimate (https://mosqlimate.org/) projects for organizing the Infodengue-Mosqlimate Sprint 2024 forecasting challenge, which motivated the development of the model presented in this paper.

## Data availability statement

The data underlying this study are publicly available at https://github. com/AlertaDengue/baseline_paper/.

## CRediT authorship contribution statement

**Laís Picinini Freitas:** Writing - original draft; Writing - review & editing; Conceptualization; Methodology; Visualization; Funding acquisition. **Danielle Andreza da Cruz Ferreira:** Writing - original draft; Writing - review & editing. **Raquel Martins Lana:** Writing - original draft; Writing - review & editing. **Daniel Cardoso Portela Câmara:** Writing - original draft; Writing - review & editing. **Tatiana P. Portella:** Writing - original draft; Writing - review & editing. **Marilia Sá Carvalho:** Writing - review & editing; Conceptualization; Funding acquisition. **Ayrton Sena Gouveia:** Writing - review & editing. **Iasmim Ferreira de Almeida:** Writing - review & editing. **Eduardo Correa Araujo:** Writing - review & editing. **Luã Bida Vacaro:** Writing - review & editing. **Fabiana Ganem:** Writing - review & editing; Funding acquisition;. **Oswaldo Gonçalves Cruz:** Writing - review & editing; Conceptualization. **Flávio Codeço Coelho:** Writing - review & editing; Conceptualization. **Claudia Torres Codeço:** Writing - review & editing; Conceptualization. **Luiz Max Carvalho:** Writing - original draft; Methodology; Writing - review & editing. **Leonardo Soares Bastos:** Writing - original draft; Writing - review & editing; Conceptualization; Methodology; Software; Data curation; Formal analysis; Funding acquisition; Project administration.

## Appendix A. Supplementary Figures

**Figure A.1:**
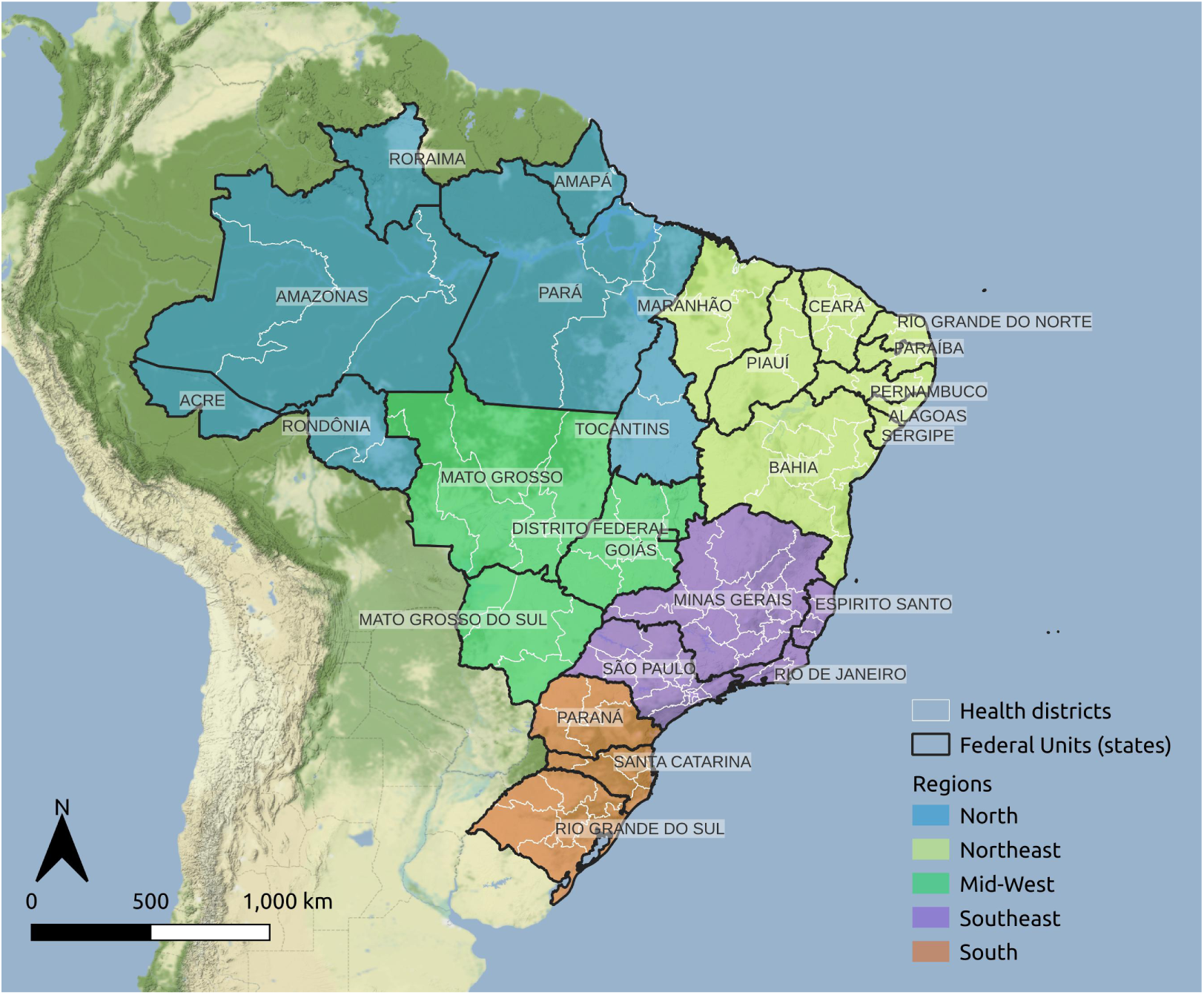
Divisions of Brazil’s territory, 2020.

**Figure A.2:**
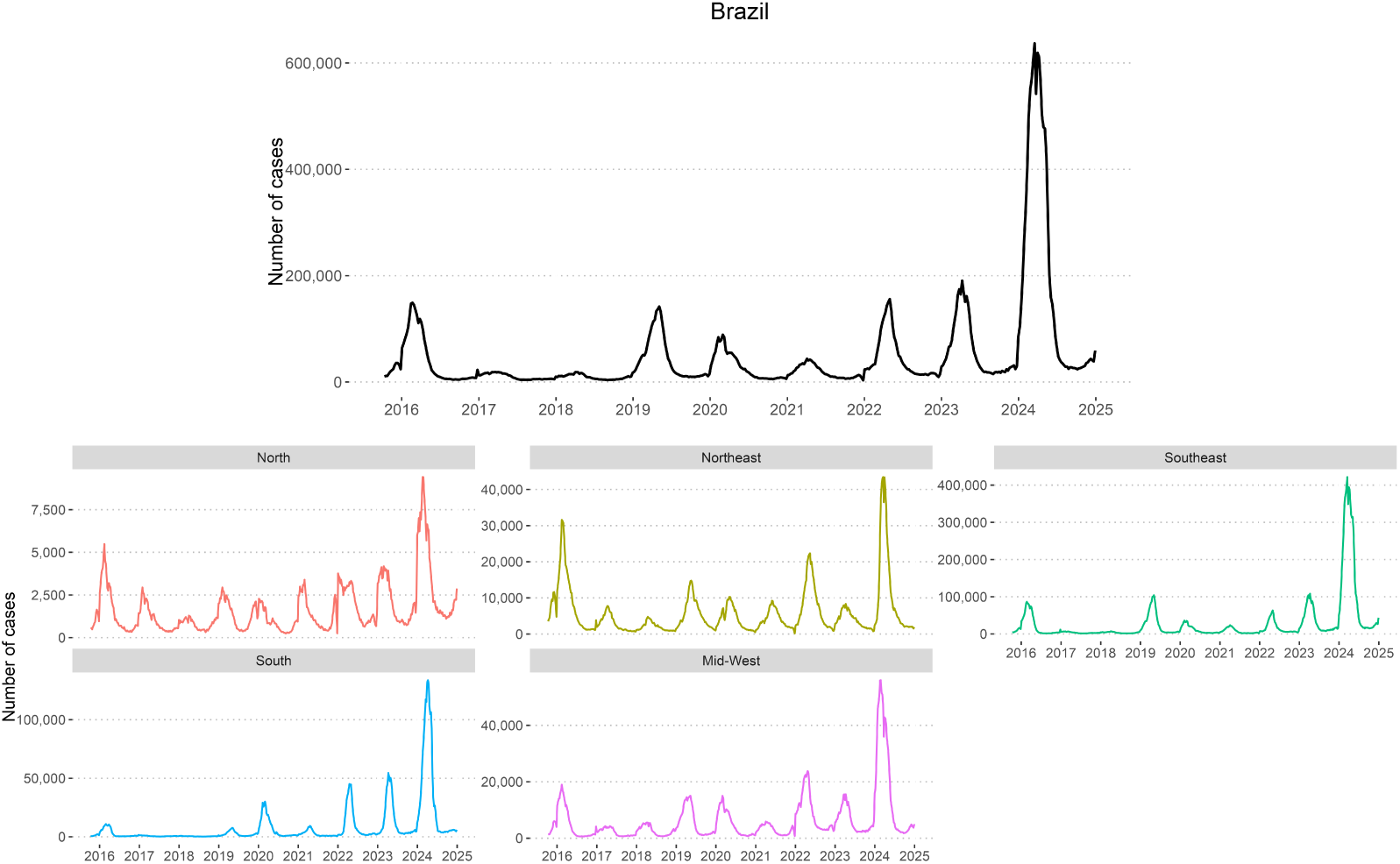
Weekly number of probable dengue cases in Brazil and by region, 2015-2024.

